# A Simulation Study Comparing Multiple Imputation and Complete Case Analysis for Handling Missing Preschool Body Mass Index

**DOI:** 10.64898/2026.08.13.26360115

**Authors:** Anamaria Savu, Douglas C. Dover, Morteza Hajihosseini, Lindsay Gaudet, Padma Kaul

## Abstract

**Introduction:** Missing data frequently occurs in health databases and can bias analyses if not correctly dealt with.

**Objectives:** Using real-world data where missing values were present in as much as 30% of our sample, we compared complete-case and multiple-imputation methods for recovering true parameters of a multivariable logistic regression model for the association between maternal glucose levels during pregnancy and child excess weight at preschool age.

**Methods:** This study utilized a cohort of 130,424 children from two Canadian urban health zones with complete preschool-age body mass index (BMI) measurements linked to administrative health data. We introduced missingness through deletion following three distinct mechanisms: missing completely at random (MCAR), at random (MAR), and not at random (MNAR). We employed complete-case and multiple-imputation methods to handle the introduced missingness. The associations between five categories of maternal glucose levels during pregnancy with child excess weight at pre-school age were determined from logistic regression models using the full observed data (true values), observed data without deletions (complete-case estimates), and imputed data (multiple-imputation estimates). Accuracy of complete-case and multiple-imputation estimates were evaluated against true values. Finally, we conducted a sensitivity analysis for the MNAR mechanism using pattern-mixture models with an additive shift.

**Results:** Under MCAR and MAR, multiple-imputation generally yielded larger bias and relative bias, but smaller or similar mean square error and reached higher significance than complete-case analysis. Both methods showed high significance (≥ 0.96) for most effects and high coverage (≥ 0.99) consistently. Under MNAR, both complete-case and multiple-imputation showed poor performance regarding bias and statistical significance. Sensitivity analysis indicated performance varied by specific effect.

**Conclusions:** When faced with missing data, researchers should assess missingness mechanisms, report both complete-case and multiple-imputation estimates under MCAR/MAR while accounting for power-versus-bias tradeoffs, and employ pattern-mixture sensitivity analyses to test robustness when MNAR is plausible.

## INTRODUCTION

Missing data represents a pervasive challenge in health databases, potentially compromising the validity and power of statistical analyses. Two common strategies for addressing missing data are complete-case analysis and multiple-imputation. Complete-case analysis offers a straightforward approach, both conceptually and practically, by excluding any observations with missing values. However, this simplicity comes at a cost: complete-case analysis has been widely criticized for its potential to introduce bias and reduce statistical power due to the exclusion of a subset of the data.[1, 2] The possibility of bias is particularly concerning when the probability that a value is missing is related to the missing value itself or to other unobserved variables. In such scenarios, the remaining complete cases may not constitute a representative sample of the overall population.

In contrast, multiple-imputation offers a more sophisticated approach. While computationally more intensive, multiple-imputation fills in missing data. This process involves creating multiple complete datasets, each with different plausible imputations for the missing values. These datasets are then analyzed separately, and the results are subsequently pooled using specific rules to account for uncertainty inherent in the imputation process. Rubin’s seminal work laid the foundational principles for multiple-imputation, illustrating its application through various examples.[3] By leveraging information contained within incomplete cases and incorporating auxiliary variables, multiple-imputation has the potential to mitigate bias and/or enhance the precision of statistical estimates.[4, 5] Despite its relative strengths, a recent systematic review highlighted the continued dominance of complete-case analysis, employed in 53% of studies reporting missing data, while multiple-imputation was utilized in only 28%.[6]

Body mass index (BMI) data is frequently missing in health datasets.[7, 8] Specifically, BMI is missing for approximately 30% of our sample of Albertan preschool-aged children.[9] The objective of this study was to evaluate the performance of logistic regression estimates when accounting for missing data using multiple-imputation versus complete-case analysis. Specifically, we assessed the association between maternal glucose levels during pregnancy and excess weight in preschool-aged children, a binary outcome defined according to World Health Organization (WHO) criteria applied to continuous BMI.[10] To systematically evaluate estimator performance, true values were known across the full dataset prior to artificially introducing missingness.

## METHODS

### Participants

We utilized a longitudinal pregnancy-birth cohort previously described.[11–13] This Alberta cohort comprises comprehensive health data on mothers and their children, including hospitalizations, outpatient and physician visits, prescriptions, laboratory results, provincial health registry, and vital statistics. It is further linked to the well-child services database, providing child height and weight measurements at preschool immunization visits, which typically occur between the child’s 4^th^ and 7^th^ birthday.

This study included children born following singletons gestations within the catchment areas of Calgary and Edmonton Health Zones of the province of Alberta, between October 1^st^ , 2008 and March 1^st^, 2015. Children were required to reside in these health zones during their preschool years to be eligible for well-child services at public health centers in those zones. Follow-up continued until March 2022. Complete set of exclusions applied to ensure data accuracy and completeness are presented in the study flow chart (**Figure 1**).[11]

**Figure 1.**
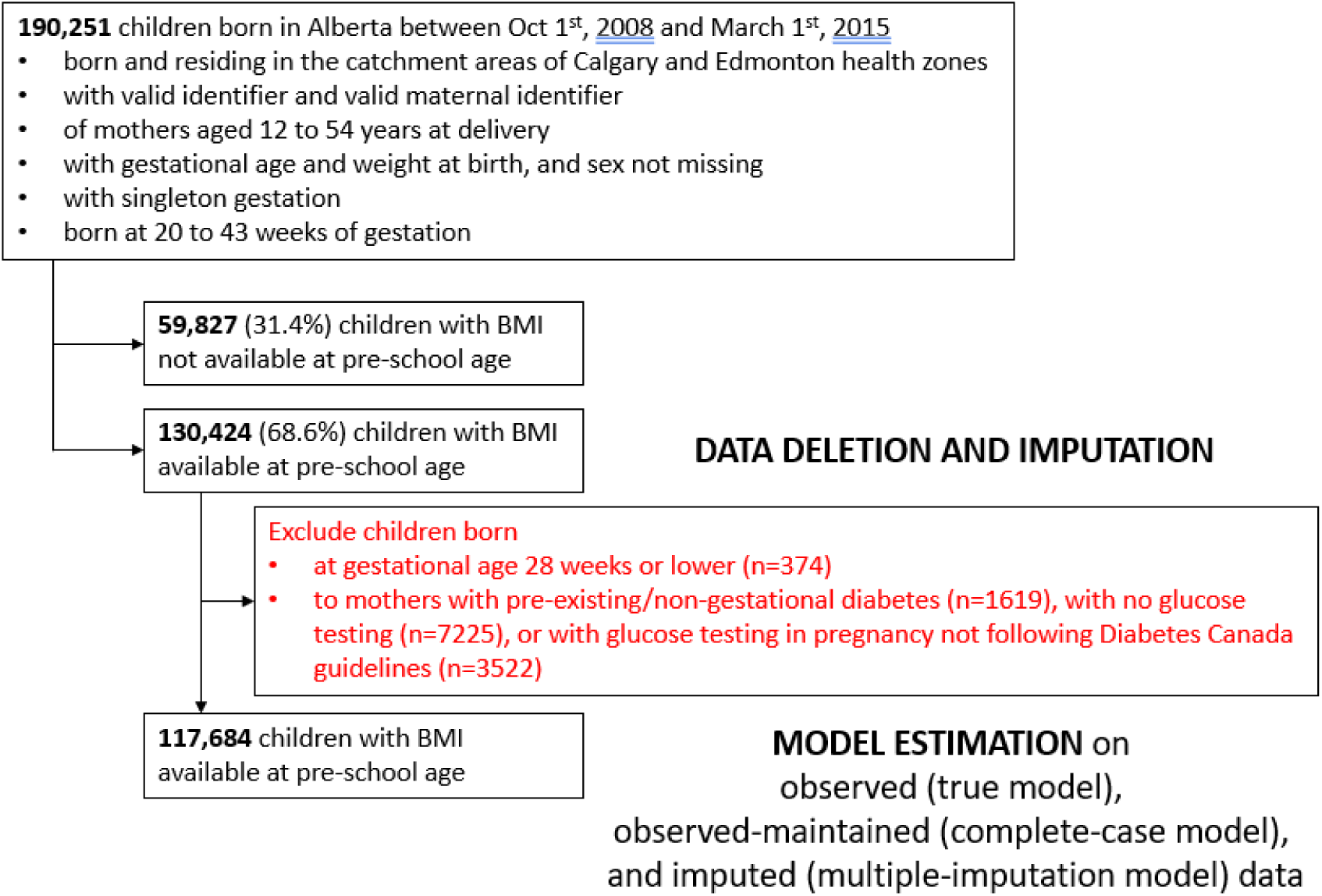
Flow chart for the selection of the study cohort. BMI = body mass index.

### Variables

#### Demographic variables

including mother age, health zone, and child sex were extracted from the provincial health care registry. Mother marital status, parity, child weigh and gestational age were determined based on vital statistics. Previously validated naming algorithms were used to identify mothers of Chinese and South Asian ethnicities.[14] The quintiles of the Canadian Index of Multiple Deprivation derived from the 2016 Statistics Canada Census data for the neighborhood of the mother were used as a measure of socioeconomic status.[15] The time of assessment of these variables was the child’s birth.

#### Comorbidities

Hypertension (pre-existing, gestational, pre-eclampsia, and eclampsia), diabetes (pre-existing, gestational), pre-existing heart failure, cardiac dysrhythmia, coronary artery disease, cerebrovascular disease, peripheral artery disease, cardiomyopathy, renal disease, and congenital heart disease were identified using established algorithms[16, 17] based on the International Classification of Diseases 10^th^ revision codes in hospitalizations, emergency department, and outpatient clinic records prior and during pregnancy. Listed comorbidities, apart from hypertension and diabetes, were combined into a no/yes indicator of cardiovascular and/or renal disease. Pharmaceutical dispenses of insulin and metformin were accessed for women who received pharmacological treatment for gestational diabetes (GDM) during pregnancy. Procedure codes on the delivery hospitalization record were used to identify children born via c-section.

#### Maternal glycaemia in pregnancy

We used two categorizations of maternal glycemia in pregnancy. The first categorization, used to impute missing BMI at preschool age, was based mainly on the GCT result: pre-existing/non-gestational diabetes prior to pregnancy, no GCT testing during pregnancy despite no pre-existing diabetes diagnosis, GCT < 7.8 mmol/L, GCT 7.8 – 11.0 mmol/L, GCT ≥ 11.1 mmol/L. The second categorization, used as an explanatory factor for a child having excess weight at pre-school age, consisted of: negative GCT (GCT < 7.8 mmol/L without collected OGTT), negative OGTT, elevated post-load glucose (PLG) alone, elevated fasting plasma glucose (FPG) only, and combined (elevated FPG with elevated PLG, either 1-hour, 2-hour, or both).[12, 13] These stem from the Diabetes Canada Clinical Practice Guidelines which recommend that all pregnant women without pre-existing diabetes are screened between 24 and 28 weeks of gestation for GDM using a two-step process of a 50-g glucose challenge test (GCT) followed by a 75-g oral glucose tolerance test (OGTT) in those with a 1-h glucose of 7.8–11.0 mmol/L on the GCT.[18] Women are identified as having GDM if their GCT is ≥ 11.1 mmol/L or if their OGTT is ≥ 5.3 mmol/L at fasting, ≥ 10.6 mmol/L at 1 h or ≥ 9.0 mmol/L at 2 h.[18]

#### Body Mass Index at Preschool Age

BMI was calculated as weight in kg/[height in m]^2^ using weight and height measurements from well-child visits. The last available measurement was used for each child. BMI z-scores were calculated using the Lambda-Mu-Sigma (LMS) method[19] and age-sex-specific parameters published by the WHO.[10] We identified children with excess weight based on WHO criteria for children aged between 5-19 years, namely BMI z score ≥ 1.[10]

### Statistical Analysis

To gain insight into the BMI missing mechanism in our data and guide our simulation of missing data we presented and compared maternal and child characteristics at birth and at preschool age between children with and without observed BMI: categorical variables using counts, percentages, and χ2 test and continuous variables using means, standard deviations (SD), and one-way ANOVA.

#### Creation of missing data

We selected the cohort of children with observed BMI data and artificially introduced missingness according to three mechanisms: missing completely at random (MCAR), missing at random (MAR), and missing not at random (MNAR). For each child, a random number, , was generated uniformly from [0, 1], and the child’s BMI value was deleted if satisfied specific conditions. This split the observed BMI data into observed-maintained and observed-deleted data. Repeating this process 100 times, yielded 100 missingness datasets per scenario.

MCAR scenario was achieved by deleting a child’s BMI if 0.3145 and maintaining, otherwise. This produced a missing rate of approximately 0.3145, aligning with the observed proportion of true missing BMI values in our dataset.[20]

MAR scenario was achieved by deleting a child’s BMI if

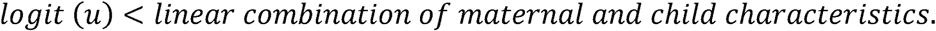

Thus, missingness in BMI depended solely on variables typically available within administrative databases, rather than on the child’s underlying BMI.[20, 21]

Based on factors previously shown to differ between children with and without available BMI data,[11] we selected maternal demographics and comorbidities and child characteristics at birth strongly correlated with excess weight. Hospital delivery status was specifically included to account for high-risk gestations and healthcare-seeking behavior, given its strong association with obstetric and early-life interventions. The coefficients for the linear combinations were found by fitting a logistic regression model for the probability of missing BMI using the cohort containing both observed and missing BMI values (**eTable 1** in the Supplement).

MNAR scenario was achieved by deleting a child’s BMI if

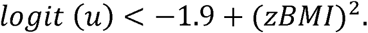

Thus, missingness in BMI was modeled as a function of the underlying BMI z-score (zBMI).

#### Imputation of created missing data

For each scenario and artificially deleted dataset we imputed the deleted BMI z score 30 times [22] using the predictive mean matching imputation method.[23, 24] Despite creating the missing data through various mechanisms, the imputation was carried under the MAR assumption using variables highly correlated with BMI or early-life interventions and maternal characteristics that were shown to be different between children with and without BMI.[11] These variables are typically available in administrative databases and are those included in **eTable 1**. Slight formatting changes were applied: birth weight (continuous z-score), preterm birth (continuous gestational age), parity (nulliparous vs. multiparous), and maternal glucose (no diabetes, pre-existing, or gestational diabetes). For each missing mechanism, we presented descriptive statistics, histograms and box plots comparing the distributions of observed-maintained, observed-deleted, and imputed BMI z scores. Descriptive statistics and histograms are based on a single imputation whereas the box plots show several imputations.

#### Maternal glucose in pregnancy and child excess weight

The model that we selected to test under our various simulated scenarios regressed the binary outcome of excess weight in the child on maternal glucose level during pregnancy and other variables: child sex, maternal demographics and comorbidities (**eTable 2** in the Supplement). Generalized estimating equation (GEE) was added to the logistic regression as it addresses the correlation between pregnancies of the same mom. This was carried within the cohort of children born at 29 or more weeks of gestation, to mothers not affected by pre-existing diabetes and underwent glucose testing according to Diabetes Canada Guidelines.[18] The odds ratios for the association between maternal glycaemia in pregnancy and child excess weight at pre-school age were estimated using the entire observed, imputed and observed-maintained BMI data. We refer to the model and associations derived based on the entire observed BMI data as the “true” model and parameter values. We refer to the model and associations estimated based on the imputed BMI data as the multiple-imputation model and estimates. Associations were estimated in each imputed data set and then combined to recover the appropriate sampling variance of the multiple-imputation estimate.[1, 3] Lastly, we refer to the model and associations estimated based on the observed-maintained BMI data as the complete-case model and estimates.

#### Multiple-imputation versus Complete-case

For each mechanism (MCAR, MAR, MNAR) and each analysis (multiple-imputation and complete-case) we identified the bias, relative bias, mean squared error, significance level, and coverage probability relative to “true” analysis across the 100 simulations. Bias was calculated as the average of the difference between estimated and “true” odds ratios. Relative bias was calculated as the average of the proportion of the difference between estimated and “true” odds ratios relative to “true” odds ratio. Mean squared error was calculated as the average of the squared difference between estimated and “true” odds ratios. Significance level was the proportion of simulations that led to a two-sided p-value < 0.05. Coverage probability was the proportion of simulations where the estimated 95% confidence interval included the “true” odds ratio.

#### Additional analysis for MNAR mechanism

Although missing data were introduced via MNAR mechanism, multiple-imputation was conducted under the MAR assumption; pattern-mixture models were therefore utilized as a sensitivity analysis. To evaluate model robustness across against deviations from the MAR assumption, an additive shift parameter ranging across clinically plausible values (−2 to 2) was applied to the imputed continuous BMI -scores. Following this adjustment, the estimated association between maternal gestational glycemia and preschool child excess weight was compared against the “true” odds ratios in terms of bias, mean squared error, statistical significance, and coverage probability.

## RESULTS

There were 190,251 children born of singleton pregnancies between 20 and 43 weeks of gestation, within the Calgary and Edmonton Health Zones, between October 1, 2008, and March 1, 2015. Among these, 130,424 children (68.6%) had height and weight data at preschool age and could have their BMI calculated.

### Missing mechanism in raw data

We presented descriptive statistics on children with and without BMI at preschool age in **eTable 3** in the Supplement. Unadjusted and adjusted analyses show differences between children with and without BMI and suggest that missingness of BMI is not MCAR.

### Imputed data

The cohort consisting of 130,424 children with BMI constituted the base of our performance assessment. For each missing mechanism we selected one round of data deletion and up to 5 instances of imputation of deleted data. **eTable 4** (Supplement) shows descriptive statistics on observed-maintained (complete-case), observed-deleted, and imputed data for the 3 mechanisms. Histograms and box-plots present further comparisons of these 3 datasets (**eFigure 1**). The zBMI distribution in the observed-maintained (complete-case), observed-deleted, and imputed data appear similar for MCAR and MAR mechanisms. The presence of numerous outliers in the MCAR and MAR box plots is a consequence of the large sample size and the typical heavy-tailed distribution of childhood BMI z-scores. However, for MNAR, the observed-deleted data exhibits substantially higher variance and more extreme values than the maintained data. Moreover, the imputed data resembles the observed-maintained (complete-case), rather than the observed-deleted data in terms of mean, outliers or density.

### True parameter values

After excluding children born at gestational age ≤ 28 weeks (n=374), of mothers without type and extent of glucose elevation, specifically of mothers affected by pre-existing diabetes (n=1,619), and of mothers who did not undergo glucose testing (n=7,225) or underwent a GCT 7.8-11.0 mmol/L or ≥11.1 mmol/L but no subsequent OGTT (n=2,379 and n=1,143) we calculated the strength of the association between maternal glucose elevation and excess weight at preschool age in children with observed BMI data (n=117,674 children of 95,346 mothers) to be: 1.09 for normal OGTT, 1.09 for elevated PGPL, 1.38 for elevated FPG, and 1.52 for combined elevation versus normal GCT after taking into account the correlation between children of the same mother (**Table1** and **eTable 2** in the Supplement).

### Bias and Relative Bias

**Table 1** shows in addition to true parameter values, the estimated values using multiple-imputation and complete-case analysis under the 3 missing mechanisms. Under MCAR and MAR, both multiple-imputation and complete-case generally exhibit relatively low bias and relative bias across most effects. However, neither method is consistently superior, as the direction and magnitude of bias vary. Compared to complete-case, multiple-imputation showed larger bias and relative bias for three effects, but lower bias for elevated FPG. The relative bias for multiple-imputation ranged from 0.5% to 2.2% under MCAR and 0.6% to 3.5% under MAR, while for complete-case, it ranged from 0.01% to 1.4% under MCAR and 0.2% to 2.1% under MAR.

**Table 1.** True and estimated effect values.

| Effect | TRUE | MCAR |  | MAR |  | MNAR |  |  |  |  |  |
| --- | --- | --- | --- | --- | --- | --- | --- | --- | --- | --- | --- |
|  |  | MI | CC | MI | CC | MI (shift = -2) | MI (shift = -1) | MI | MI (shift = 1) | MI (shift = 2) | CC |
| Normal OGTT | 1.09 | 1.08 | 1.09 | 1.08 | 1.09 | 1.01 | 1.01 | 1.04 | 1.06 | 1.05 | 1.03 |
| Elevated PLPG | 1.09 | 1.11 | 1.09 | 1.11 | 1.09 | 0.92 | 0.92 | 0.96 | 1.02 | 1.05 | 0.94 |
| Elevated FPG | 1.38 | 1.39 | 1.40 | 1.39 | 1.41 | 1.02 | 1.02 | 1.13 | 1.24 | 1.23 | 1.10 |
| Combined | 1.52 | 1.49 | 1.53 | 1.47 | 1.50 | 1.04 | 1.05 | 1.18 | 1.31 | 1.32 | 1.17 |
| Note: CC = complete case analysis; FPG = fasting plasma glucose; MAR = missing at random; MCAR = missing completely at random; MNAR = missing not at random; MI = multiple imputation; OGTT = 75-g oral glucose tolerance test; PLPG = post-load plasma glucose. |  |  |  |  |  |  |  |  |  |  |  |

Under MNAR, both multiple-imputation and complete-case primarily underestimated and showed negative bias, with complete-case generally exhibiting a larger magnitude of negative bias for all four effects. As the effect’s “true” value increases, both bias and relative bias tend to increase. For normal OGTT, the relative bias is noticeable at 4.5% (multiple-imputation) and 5.0% (complete-case), increasing substantially for combined, to 22.5% (multiple-imputation) and 23.3% (complete-case). Compared to MCAR and MAR mechanisms, the MNAR scenario substantially increases bias for both multiple imputation and complete-case analysis. This indicates that both methods struggle significantly with more complex missing data mechanisms. (**Table 2**).

**Table 2.** Performance of multiple-imputation and complete-case analysis versus “true” analysis.

| Effect | MCAR |  | MAR |  | MNAR |  |
| --- | --- | --- | --- | --- | --- | --- |
|  | MI | CC | MI | CC | MI | CC |
| **BIAS** |  |  |  |  |  |  |
| Normal OGTT | -0.008910 | 0.000158 | -0.006660 | 0.002585 | -0.048810 | -0.054920 |
| Elevated PLPG | 0.018474 | -0.002160 | 0.015625 | -0.002260 | -0.132970 | -0.151940 |
| Elevated FPG | 0.006702 | 0.018782 | 0.011187 | 0.028658 | -0.252150 | -0.276900 |
| Combined | -0.033360 | 0.007331 | -0.053800 | -0.021890 | -0.341880 | -0.354520 |
| **RELATIVE BIAS** |  |  |  |  |  |  |
| Normal OGTT | -0.008170 | 0.000145 | -0.006110 | 0.002372 | -0.044780 | -0.050390 |
| Elevated PLPG | 0.016949 | -0.001980 | 0.014335 | -0.002070 | -0.121990 | -0.139390 |
| Elevated FPG | 0.004857 | 0.013610 | 0.008107 | 0.020767 | -0.182720 | -0.200650 |
| Combined | -0.021950 | 0.004823 | -0.035390 | -0.014400 | -0.224920 | -0.233240 |
| **MEAN SQUARED ERROR** |  |  |  |  |  |  |
| Normal OGTT | 0.000191 | 0.000210 | 0.000134 | 0.000179 | 0.002598 | 0.003430 |
| Elevated PLPG | 0.000759 | 0.000748 | 0.000585 | 0.000565 | 0.018342 | 0.024376 |
| Elevated FPG | 0.003749 | 0.008360 | 0.004689 | 0.009878 | 0.070552 | 0.091811 |
| Combined | 0.003713 | 0.004979 | 0.005208 | 0.004875 | 0.120925 | 0.135267 |
| **COVERAGE** |  |  |  |  |  |  |
| Normal OGTT | 1.00 | 0.99 | 1.00 | 1.00 | 0.76 | 0.61 |
| Elevated PLPG | 1.00 | 1.00 | 1.00 | 1.00 | 0.33 | 0.22 |
| Elevated FPG | 1.00 | 1.00 | 1.00 | 1.00 | 0.93 | 0.78 |
| Combined | 1.00 | 1.00 | 1.00 | 1.00 | 0.43 | 0.46 |
| **STATISTIAL SIGNIFICANCE** |  |  |  |  |  |  |
| Normal OGTT | 1.00 | 1.00 | 1.00 | 1.00 | 0.09 | 0.10 |
| Elevated PLPG | 0.75 | 0.40 | 0.79 | 0.40 | 0.00 | 0.08 |
| Elevated FPG | 0.98 | 0.96 | 0.99 | 0.96 | 0.04 | 0.07 |
| Combined | 1.00 | 1.00 | 1.00 | 1.00 | 0.05 | 0.08 |
| Note: CC = complete case analysis; FPG = fasting plasma glucose; MAR = missing at random; MCAR = missing completely at random; MNAR = missing not at random; MI = multiple imputation analysis; OGTT = 75-g oral glucose tolerance test; PLPG = post-load plasma glucose. Effects of maternal glucose level in pregnancy are relative to normal 50-g glucose challenge test. |  |  |  |  |  |  |

### Mean squared error

Under MCAR and MAR scenarios, the mean squared errors of the estimates from multiple-imputation and complete-case were comparable for 3 of the 4 effects (normal OGTT, elevated PLPG, and Combined), with no clear pattern of superiority for either method. Notably, for elevated FPG, multiple-imputation versus complete-case displayed mean squared error width reduction: 55% under MCAR and 52% under MAR. Under MNAR, multiple-imputation consistently resulted in slightly lower square errors across all four effects, possibly due to multiple-imputation picking up some residual correlation with unmeasured effects (**Table 2**).

### Coverage

Coverage probability ranged from 99% to 100% for all four effects under MCAR and MAR, regardless of the imputation method. However, under MNAR, coverage probability varied: it was high for elevated FPG (93% for multiple-imputation and 78% for complete-case), moderate for normal OGTT (76% for multiple-imputation and 61% for complete-case), and low for the remaining two effects (33-43% for multiple-imputation and 22-46% for complete-case).

For three out of the four effects, multiple-imputation exhibited higher coverage probability than complete-case (**Table 2**).

### Significance

Statistical power for detecting normal OGTT, elevated FPG, and combined elevations (using both multiple-imputation and complete-case methods) was high, ranging from 96% to 100% under both MCAR and MAR missing data mechanisms. However, for the effect of elevated PLPG, multiple-imputation demonstrated improved power compared to complete-case, restoring between 35% and 39% of the lost power: 75% versus 40% under MCAR, and 79% versus 40% under MAR. Notably, under MNAR, the power of both multiple-imputation and complete-case were similarly low and unacceptable, ranging from 0% to 9%. This indicates that under MNAR, both multiple-imputation and complete-case analyses are unlikely to detect a true effect, in our specific example underestimating the true effect (**Table 2**).

### Sensitivity analysis of MNAR data

**Figure 2** and **Table 1** illustrate the performance of the pattern-mixture model in analyzing MNAR data. To assess the model’s ability to recover true values under the MNAR mechanism, we varied the additive shift across clinically plausible values: -2 and 2. For all effects, absolute value of bias and relative bias, and mean squared error decreased as the shift increased, reaching a threshold around a shift value of 1 beyond which minimal improvements were observed. These thresholds were dependent on the effect size, with larger effects exhibiting larger thresholds and smaller effects exhibiting smaller thresholds. Underestimation was consistent across all effects and shift values. Coverage generally increased with the shift, reaching a peak before decreasing for three effects (normal OGTT, elevated FPG, and combined) or plateauing for elevated PLPG. The peak coverage was above 0.9 for normal OGTT (at a shift between 0.2 and 1.5), elevated FPG (at a shift between 0.3 and 1.2), and elevated PLPG (at a shift between 1 and 2), and 0.69 for the combined effect (at a shift of 0.7). Significance displayed a distinct pattern. For three effects it started near 0.7 at the lowest shift value (−2), it decreased to below 0.1 before increasing for positive shift values. Significance reached a threshold above 0.9 for normal OGTT and the combined effect, just above 0.7 for elevated FPG. For elevated PLPG it started near 0.7 at the lowest shift value, kept decreasing afterwards reaching a value close to 0, and starting a mild increase at a shift value of around 0.5.

**Figure 2.**
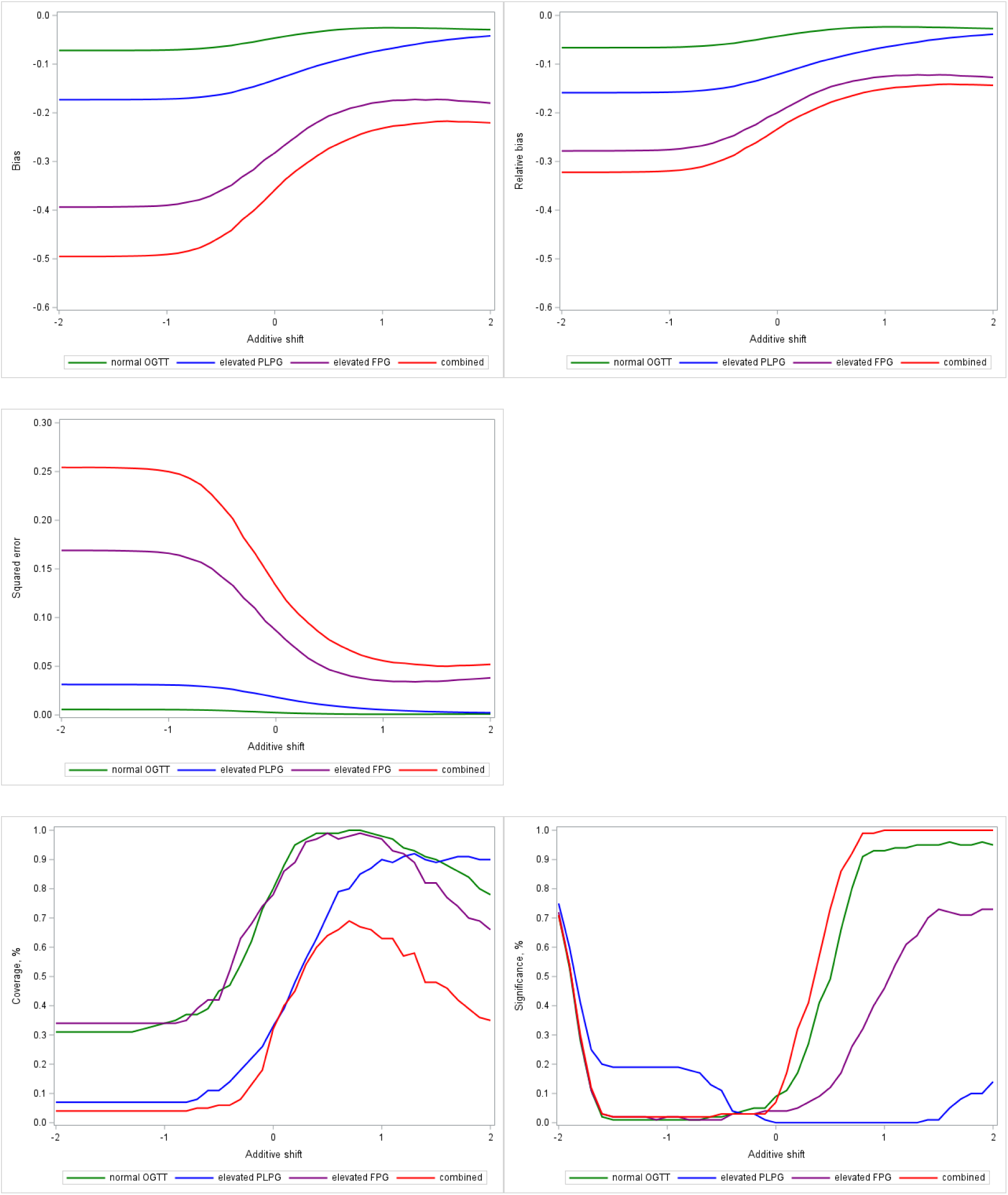
Performance of pattern-mixture model under MNAR. Note: FPG = fasting plasma glucose; MNAR = missing not at random; OGTT = 75-g oral glucose tolerance test; PLPG = post-load plasma glucose. Effects of maternal glucose level in pregnancy are relative to normal 50-g glucose challenge test.

In summary, statistical inference varied substantially across shift values. For instance, the effect of combined, the effect with the largest magnitude 1.52, was estimated as 1.05 and statistically non-significant under a shift value of −1, representing that missing BMI values are on average 1 standard deviation below observed cases. With no shift, the estimate increased to 1.18 but remained non-significant. Moving the shift to +1 raised the estimate to 1.31 and displayed statistical significance. This sensitivity in both point estimates and significance levels is characteristic of MNAR data, providing empirical validation of the simulated MNAR mechanism.

## DISCUSSION

This study compared the performance of multiple-imputation using predictive mean matching against complete-case analysis for estimating a multivariable logistic model via generalized estimating equations. The data, where the “true model” parameters were known due to complete outcome data, underwent simulated missingness under three scenarios: MCAR, MAR, and MNAR. The outcome of interest was excess weight at preschool age (BMI z-score ≥ 1), a variable often with missing values in health databases.[11]

Under MCAR and MAR, multiple-imputation showed larger bias and relative bias for three of the four estimated effects compared to CC. This bias may reflect minor misspecifications in the imputation model’s functional form or the inclusion of auxiliary variables that added more noise than predictive value. However, multiple-imputation yielded a gain in power for one effect and similar power for the other three, demonstrating its ability to recover statistical power by preserving the full sample size and reducing standard errors. Estimators with lower mean squared error are preferred because they yield estimates closer to the true parameter value. When mean squared error values are comparable, the estimator with lower bias is favored. Based on this criterion, multiple-imputation is preferred over complete-case analysis under MCAR for all four effects, and under MAR for three of the four effects, the exception being the effect with the largest magnitude.

Under MNAR, both multiple-imputation and complete-case demonstrated unsatisfactory performance, as previously observed.[25, 26] Pattern-mixture modelling under MNAR assumption showed that the performances depend on the specific effect, with an acceptable shift value found for only one effect. This variability highlights the challenges of conducting meaningful sensitivity analyses, which rely on unverifiable assumptions about the exact shift parameter across different types of effects.

These simulations suggest that multiple-imputation may offer, for weak associations, a gain in power under MCAR and MAR despite increased bias. While multiple-imputation nor complete-case adequately addressed MNAR, the pattern-mixture model tested if the estimations changed under different realistic assumptions about the missing data.

In summary, health researchers faced with missing data first should investigate the underlying missingness mechanism. Comparing key characteristics between observations with and without missing values can be used to evaluate whether the missing data satisfies the MCAR assumption; a rejection of these tests indicating that the data is likely MAR or MNAR. Distinguishing between MAR and MNAR is challenging because statistical tests cannot evaluate unobserved values. Consequently, researchers must rely on clinical knowledge and literature reviews to assess whether an MNAR scenario is plausible. In the context of childhood obesity research, for instance, families with the highest-weight children might skip preschool BMI checks because they are under specialized physician care.

### Strengths

Firstly, we leveraged a substantial sample size derived from real-world data, enhancing the practical relevance of our findings. This large dataset allowed for a robust investigation into the performance of multiple-imputation compared to complete-case analysis. Secondly, our focus on BMI at pre-school age, a critical variable in health research with significant implications for long-term health outcomes, underscores the importance and applicability of our findings to the broader health research community. By examining a variable commonly affected by missing data in health databases, our study provides valuable insights into handling this challenge in a context of significant public health interest.

### Limitations

Despite its strengths, our study also has certain limitations that warrant consideration. Firstly, our investigation of MNAR mechanisms was limited to a single type. This specific MNAR may not fully represent the diverse and complex ways in which data can be missing in real-world scenarios. Consequently, the generalizability of our findings regarding MNAR to other studies with different underlying missing data mechanisms may be limited. Secondly, our analysis focused on only four specific effects, which spanned a limited range of values and levels of statistical significance. This restricted focus might not fully capture the nuances in performance of the multiple-imputation and complete-case methods across a wider spectrum of effect sizes and statistical evidence. It is possible that the relative advantages or disadvantages of each method could vary depending on the magnitude of the estimated effects. Future research could benefit from exploring a broader range of MNAR scenarios and a more diverse set of parameters, encompassing a broader range of effect sizes, including those with varying degrees of statistical significance and practical importance, to provide a more comprehensive understanding of the methods’ performance.

## CONCLUSION

The choice of analytical approach for handling missing data inherently depends on underlying assumptions about the missingness mechanism. If non-attendance at preschool well-child visits represents a completely random sample of the target population, the data are MCAR, making complete-case analysis appropriate. Conversely, if missing visits correlate with observed maternal or child characteristics, such as demographics or comorbidities, the data are MAR. Under MAR, multiple-imputation is required, while complete-case analysis is unsuitable and introduces bias.

When operating under plausible MCAR or MAR assumptions, researchers should present both complete-case and multiple-imputation analyses, systematically comparing their point estimates and confidence intervals. However, caution is necessary: a statistically significant result following multiple-imputation for a small effect size may reflect increased statistical power, which can be accompanied by inflated bias. If an effect gains statistical significance under multiple-imputation with minimal shift in the point estimate, the finding likely reflects enhanced power.

When missingness depends on unobserved factors, such as children with extreme BMI values missing visits because they are under specialized physician care, the data are MNAR. Because both complete-case analysis and standard multiple-imputation fail under MNAR, researchers should avoid relying on a single model and instead present a range of sensitivity analyses. In these scenarios, a pattern-mixture model sensitivity analysis is particularly valuable. Following standard multiple-imputation, the pattern-mixture shifts the imputed values for missing cases across plausible ranges. If the overall conclusions remain robust after these manual shifts, it provides evidence that the primary findings are not meaningfully distorted by potential MNAR mechanisms.

## ACKNOWLEDGEMENTS

This study is based in part on data provided by PPHS (Primary & Preventative Health Services) and Alberta Health Services. We thank the Customer Relationship Management and Data Access Unit at PPHS for creating the linked database. The interpretation and conclusions are those of the researchers and do not represent the views of the Government of Alberta. Neither the Government of Alberta nor PPHS express any opinion in relation to this study.

## Ethics Statement

This study was approved by the University of Alberta Research Ethics Board (Pro00056999). The ethics panel waived the requirement for consent after determining that the research was a retrospective database review for which participant consent for access to personally identifiable health information would not be reasonable, feasible or practical.

## Data Availability Statement

The data underlying this article was provided by the Government of Alberta under the terms of a research agreement. Inquiries regarding access to the data can be made to

## AI Disclosure Statement

During the preparation of this work, the authors used Gemini to assist with text editing and grammar refinement. The authors reviewed and edited the output as needed and take full responsibility for the content. All conceptual frameworks, structural choices, and design components remain exclusively the property of the authors.

## SUPPLEMENTARY MATERIALS

**eTable 1.** Multivariable model for the outcome of child having missing body mass index at pre-school age (n=190,251).

**eTable 2.** Multivariable model for the outcome of child having excess weight at pre-school age (n=117,684).

**eTable 3.** Descriptive statistics: Children with missing versus those with available body mass index at pre-school age.

**eTable 4.** Descriptive statistics: Observed versus imputed z-score of body mass index at preschool age by missing mechanism

**eFigure 1.** Histogram and boxplot of imputed versus observed z-score body mass index for 4 missing mechanisms

## ABBREVIATIONS

BMI: body mass index
CI: confidence interval
CVDR: cardiovascular and/or renal disease
FPG: fasting plasma glucose
GCT: glucose challenge test
GDM: gestational diabetes
GEE: generalized estimating equation
LMS: Lambda-Mu-Sigma method
MAR: missing at random
MCAR: missing completely at random
MNAR: missing not at random
OBS: observed
OGTT: 75-g oral glucose tolerance test
PLPG: post-load plasma glucose
Q: quartile or quintile
SD: standard deviation
WHO: world health organization

