## Supplementary Materials for "A Simulation Study Comparing Multiple Imputation and Complete Case Analysis for Handling Missing Preschool Body Mass Index"

### eTable 1. Multivariable model for the outcome of child having missing body mass index at pre-school age (n=190,251).

|  | **Odds Ratio** | **95%** | **CI** | **P-value** |
| --- | --- | --- | --- | --- |
| Child Male Sex | 1.03 | *(1.01,* | *1.05)* | 0.0042 |
| Birth Weight (ref: 2500 - 3999g), < 2500g | 1.06 | *(1.00,* | *1.11)* | 0.0426 |
| ≥ 4000g | 1.06 | *(1.03,* | *1.10)* | 0.0004 |
| Preterm Birth | 0.99 | *(0.95,* | *1.04)* | 0.7534 |
| Maternal Age at Birth per 5 years | 0.98 | *(0.98,* | *0.98)* | <.0001 |
| Mother Married at Birth | 0.62 | *(0.60,* | *0.63)* | <.0001 |
| Parity (ref: 0), 1 | 1.39 | *(1.36,* | *1.42)* | <.0001 |
| 2 | 1.87 | *(1.81,* | *1.93)* | <.0001 |
| ≥ 3 | 2.74 | *(2.63,* | *2.86)* | <.0001 |
| Situational Vulnerability (ref: Q1), Q2 | 1.05 | *(1.02,* | *1.08)* | 0.0004 |
| Q3 | 1.05 | *(1.02,* | *1.09)* | 0.0013 |
| Q4 | 1.15 | *(1.11,* | *1.19)* | <.0001 |
| Q5 | 1.39 | *(1.34,* | *1.44)* | <.0001 |
| Missing | 1.00 | *(0.87,* | *1.15)* | 0.9953 |
| Economic Dependency (ref: Q1), Q2 | 0.94 | *(0.92,* | *0.96)* | <.0001 |
| Q3 | 1.01 | *(0.98,* | *1.04)* | 0.4199 |
| Q4 | 0.94 | *(0.91,* | *0.98)* | 0.0008 |
| Q5 | 0.94 | *(0.90,* | *0.99)* | 0.0093 |
| Ethnicity (ref: General Population), Chinese | 0.72 | *(0.68,* | *0.76)* | <.0001 |
| South Asian | 0.70 | *(0.66,* | *0.74)* | <.0001 |
| Health Zone (ref: Calgary), Edmonton | 0.95 | *(0.93,* | *0.98)* | <.0001 |
| Maternal Glucose (ref: GCT < 7.8mmol/L), GCT: 7.8-11.0 mmol/L | 1.03 | *(1.00,* | *1.06)* | 0.0288 |
| GCT ≥ 11.1mmol/L | 1.24 | *(1.13,* | *1.37)* | <.0001 |
| Pre-existing Diabetes | 1.08 | *(0.99,* | *1.18)* | 0.0991 |
| Not Tested | 1.58 | *(1.52,* | *1.63)* | <.0001 |
| Pre-existing/Non-gestational Hypertension | 1.02 | *(0.96,* | *1.09)* | 0.5426 |
| Gestational Hypertension | 0.96 | *(0.92,* | *1.01)* | 0.0954 |
| Pre-eclampsia/Eclampsia | 1.03 | *(0.96,* | *1.11)* | 0.3620 |
| CVRD | 1.11 | *(1.05,* | *1.16)* | <.0001 |
| Hospital Birth | 0.54 | *(0.51,* | *0.57)* | <.0001 |

Note: CI: confidence interval; CVRD: cardiovascular and/or renal disease; GCT: 50g glucose challenge test; Q: quintile. Discrimination index = 0.63.

### eTable 2. Multivariable model for the outcome of child having excess weight at pre-school age (n=117,684).

|  | **Odds Ratio** | **95%** | **CI** | **P-value** |
| --- | --- | --- | --- | --- |
| Maternal Glucose in Pregnancy (ref: normal GCT) normal OGTT | 1.09 | *(1.05,* | *1.13)* | <.0001 |
| elevated PLPG | 1.09 | *(1.01,* | *1.17)* | 0.0264 |
| elevated FPG | 1.38 | *(1.16,* | *1.65)* | 0.0003 |
| combined | 1.52 | *(1.31,* | *1.76)* | <.0001 |
| Child Male Sex | 1.32 | *(1.28,* | *1.35)* | <.0001 |
| Maternal Age at Birth per 5 years | 0.99 | *(0.99,* | *1.00)* | <.0001 |
| Mother Married at Birth | 0.74 | *(0.71,* | *0.77)* | <.0001 |
| Parity (ref: 0), ≥1 | 0.99 | *(0.96,* | *1.01)* | 0.3253 |
| Economic Dependency (ref: Q1), Q2 | 1.02 | *(0.98,* | *1.06)* | 0.2950 |
| Q3 | 1.03 | *(0.99,* | *1.08)* | 0.1293 |
| Q4 | 1.00 | *(0.95,* | *1.04)* | 0.8567 |
| Q5 | 1.03 | *(0.97,* | *1.10)* | 0.3076 |
| missing | 0.85 | *(0.55,* | *1.29)* | 0.4400 |
| Ethnicity (ref: General Population), Chinese | 0.61 | *(0.56,* | *0.65)* | <.0001 |
| South Asian | 0.93 | *(0.87,* | *1.00)* | 0.0624 |
| Health Zone (ref: Calgary), Edmonton | 0.73 | *(0.71,* | *0.76)* | <.0001 |
| Pre-existing/Non-gestational Hypertension | 1.20 | *(1.10,* | *1.31)* | <.0001 |
| Gestational Hypertension | 1.22 | *(1.15,* | *1.29)* | <.0001 |
| Pre-eclampsia/Eclampsia | 1.11 | *(1.01,* | *1.22)* | 0.0378 |
| CVRD | 1.06 | *(0.98,* | *1.13)* | 0.1392 |
| Pharmacological Treatment for GDM (Metformin, Insulin) | 1.17 | *(1.05,* | *1.31)* | 0.0063 |

Note: CI: confidence interval; CVRD: cardiovascular and/or renal disease; FPG: fasting plasma glucose; GCT: 50g glucose challenge test; GDM: gestational diabetes; OGTT: 75g oral glucose tolerance test; PLPG: post-load plasma glucose; Q: quintile. Discrimination index = 0.582.

### eTable 3. Descriptive statistics: Children with missing versus those with available body mass index at pre-school age.

| **Characteristic** | **Available BMI** | **Missing BMI** | **P-value** |
| --- | --- | --- | --- |
| Total N | 130424 | 59827 |  |
| Child Sex, Female | 63541 (48.7) | 28778 (48.1) | 0.0124 |
| Male | 66883 (51.3) | 31049 (51.9) |  |
| Birth Weight, < 2500g | 6695 (5.1) | 3196 (5.3) | <.0001 |
| 2500-3999g | 112210 (86.0) | 50584 (84.6) |  |
| ≥ 4000g | 11519 (8.8) | 6047 (10.1) |  |
| Mean Gestational Age at Birth (SD) (week) | 38.9 (1.7) | 38.8 (1.8) | 0.0092 |
| Preterm Birth | 8556 (6.6) | 4141 (6.9) | 0.0034 |
| Mean Maternal Age at Birth (SD) (year) | 31.0 (5.0) | 30.3 (5.5) | <.0001 |
| Mother Married at Birth | 103906 (79.7) | 40582 (67.8) | <.0001 |
| Maternal Parity, 0 | 60457 (46.4) | 22270 (37.2) | <.0001 |
| 1 | 47386 (36.3) | 21222 (35.5) |  |
| 2 | 15844 (12.1) | 9630 (16.1) |  |
| ≥ 3 | 6737 (5.2) | 6705 (11.2) |  |
| Situational Vulnerability, Q1 (least deprived) | 42705 (32.7) | 16248 (27.2) | <.0001 |
| Q2 | 31774 (24.4) | 13377 (22.4) |  |
| Q3 | 24538 (18.8) | 11000 (18.4) |  |
| Q4 | 18440 (14.1) | 9624 (16.1) |  |
| Q5 (most deprived) | 12359 (9.5) | 9124 (15.3) |  |
| Missing | 608 (0.5) | 454 (0.8) |  |
| Economic Dependency, Q1 (least deprived) | 52009 (39.9) | 23396 (39.1) | <.0001 |
| Q2 | 35133 (26.9) | 15526 (26.0) |  |
| Q3 | 19920 (15.3) | 10136 (16.9) |  |
| Q4 | 14267 (10.9) | 6463 (10.8) |  |
| Q5 (most deprived) | 8487 (6.5) | 3852 (6.4) |  |
| Missing | 608 (0.5) | 454 (0.8) |  |
| Ethnicity, Chinese | 7394 (5.7) | 2015 (3.4) | <.0001 |
| South Asian | 5883 (4.5) | 1623 (2.7) | <.0001 |
| Health Zone, Calgary | 72224 (55.4) | 32556 (54.4) | <.0001 |
| Edmonton | 58200 (44.6) | 27271 (45.6) |  |
| Maternal Glucose, GCT < 7.8mmol/L | 93599 (71.8) | 39739 (66.4) | <.0001 |
| GCT: 7.8-11.0mmol/L | 25037 (19.2) | 10549 (17.6) |  |
| GCT ≥ 11.1mmol/L | 1299 (1.0) | 659 (1.1) |  |
| Pre-existing Diabetes | 1626 (1.2) | 782 (1.3) |  |
| Not Tested | 8863 (6.8) | 8098 (13.5) |  |
| Pre-existing/Non-gestational Hypertension | 3359 (2.6) | 1602 (2.7) | 0.1937 |
| Gestational Hypertension | 7938 (6.1) | 3201 (5.4) | <.0001 |
| Pre-eclampsia/Eclampsia | 2738 (2.1) | 1216 (2.0) | 0.3431 |
| CVRD | 5125 (3.9) | 3028 (5.1) | <.0001 |
| Hospital Birth | 128126 (98.2) | 57285 (95.8) | <.0001 |

Note: BMI: body mass index; CVRD: cardiovascular and/or renal disease; GCT: 50g glucose challenge test; Q: quintile; SD: standard deviation.

### eTable 4. Descriptive statistics: Observed versus imputed z-score of body mass index at preschool age by missing mechanism

|  | | **N Children** | | **Min** | | **Q1** | | **Median** | | **Q3** | | **Max** | | **Mean** | | **SD** |
| --- | --- | --- | --- | --- | --- | --- | --- | --- | --- | --- | --- | --- | --- | --- | --- | --- |
| MCAR | OBS-MAINTAINED | | 89,355 | | -7.9560 | | -.3581 | | 0.2492 | | 0.9085 | | 7.9712 | | 0.3410 | 1.1011 |
| MCAR | OBS-DELETED | | 41,069 | | -7.8255 | | -.3581 | | 0.2801 | | 0.9218 | | 7.7895 | | 0.3496 | 1.0886 |
| MCAR | IMPUTED | | 41,069 | | -6.4587 | | -.3918 | | 0.2294 | | 0.8842 | | 7.9712 | | 0.3031 | 1.0894 |
| MAR | OBS-MAINTAINED | | 91,132 | | -7.9560 | | -.3693 | | 0.2403 | | 0.8989 | | 7.9712 | | 0.3310 | 1.0931 |
| MAR | OBS-DELETED | | 39,292 | | -7.8255 | | -.3271 | | 0.2949 | | 0.9536 | | 7.7895 | | 0.3733 | 1.1061 |
| MAR | IMPUTED | | 39,292 | | -7.9560 | | -.3910 | | 0.2328 | | 0.8989 | | 7.9712 | | 0.3202 | 1.0971 |
| MNAR | OBS-MAINTAINED | | 90,200 | | -2.7317 | | -.3667 | | 0.1518 | | 0.6310 | | 2.7711 | | 0.1349 | 0.7042 |
| MNAR | OBS-DELETED | | 40,224 | | -7.9560 | | -.3283 | | 0.9182 | | 1.8197 | | 7.9712 | | 0.8120 | 1.5729 |
| MNAR | IMPUTED | | 40,224 | | -2.2530 | | -.3912 | | 0.1053 | | 0.6128 | | 2.6298 | | 0.1120 | 0.7147 |

Note: MAR: missing at random; MCAR: missing completely at random; MNAR: missing not at random; OBS: observed; Q: quartile; SD: standard deviation.

### eFigure 1. Histogram and boxplot of imputed versus observed z-score body mass index for 4 missing mechanisms

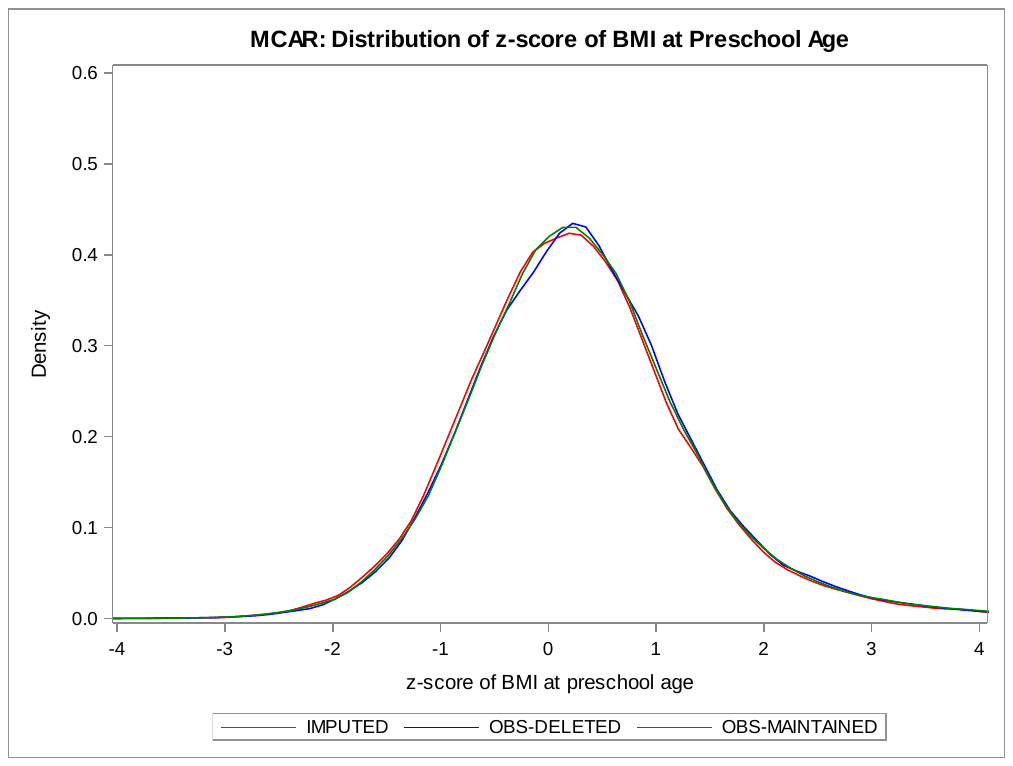

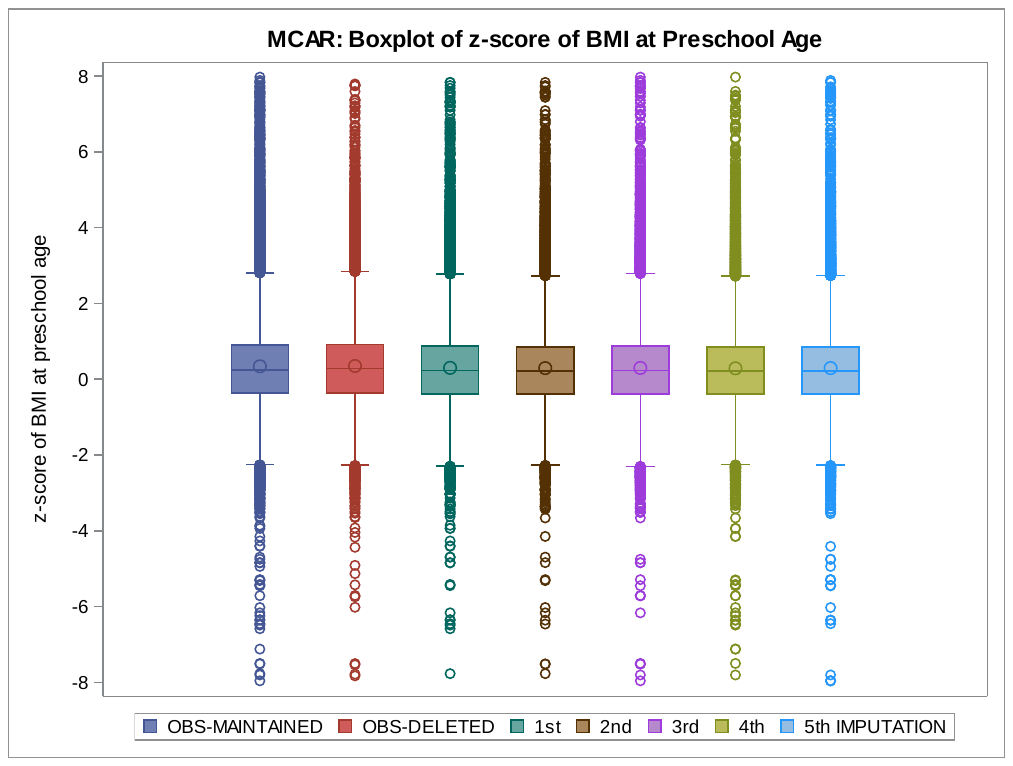

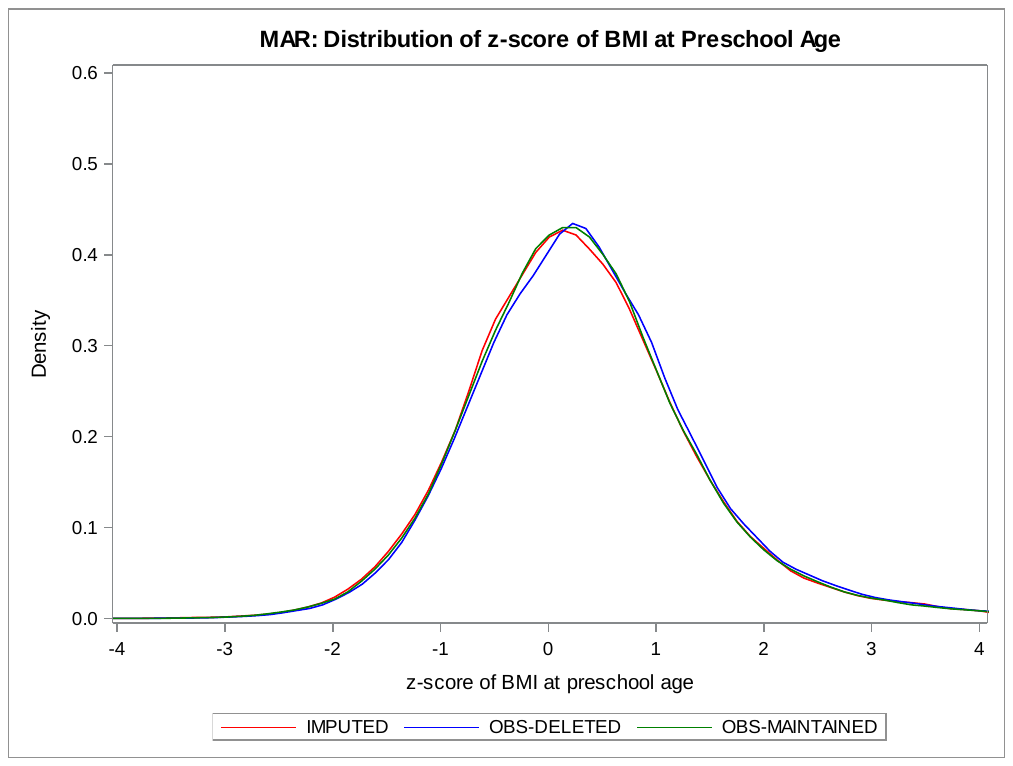

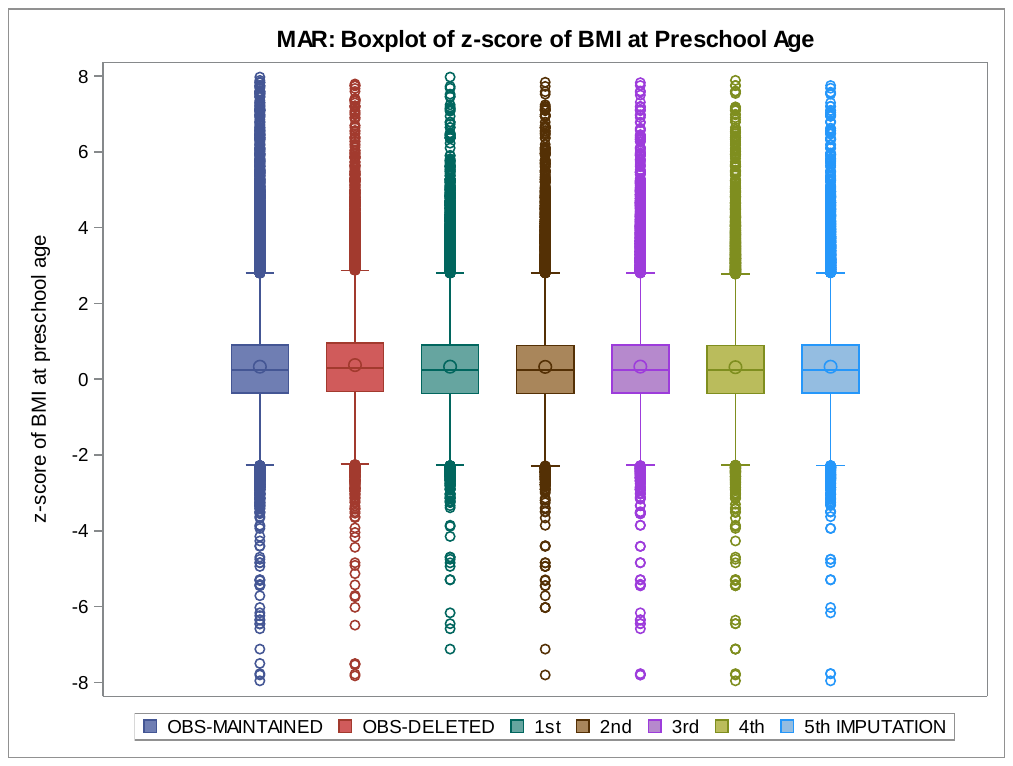

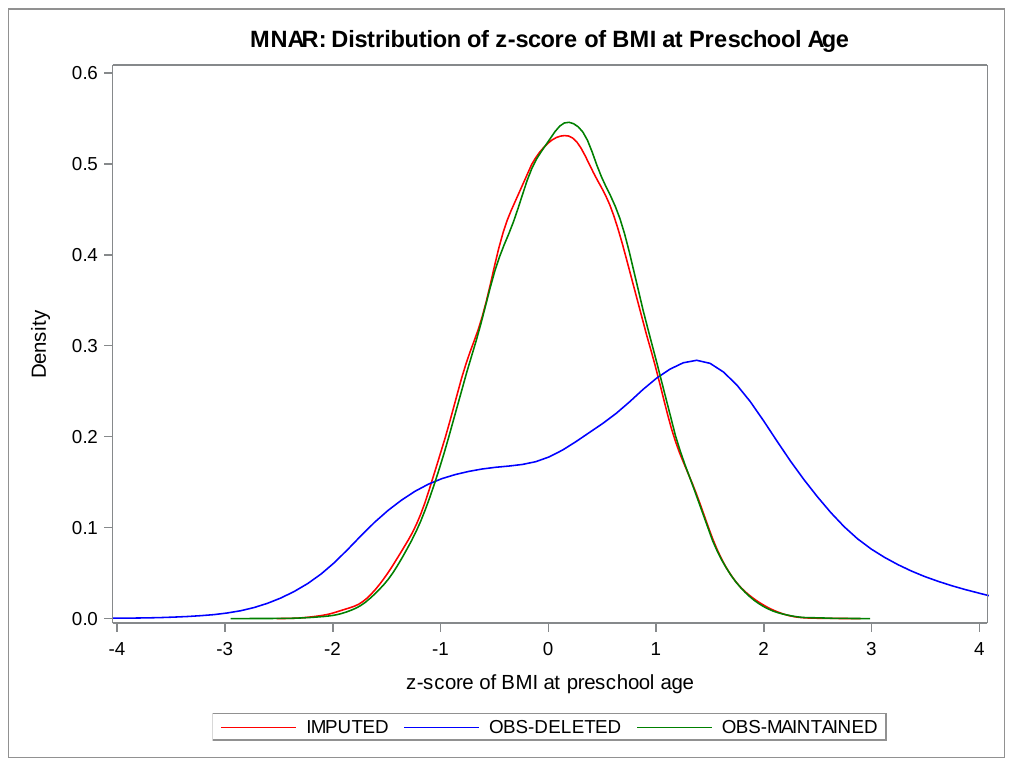

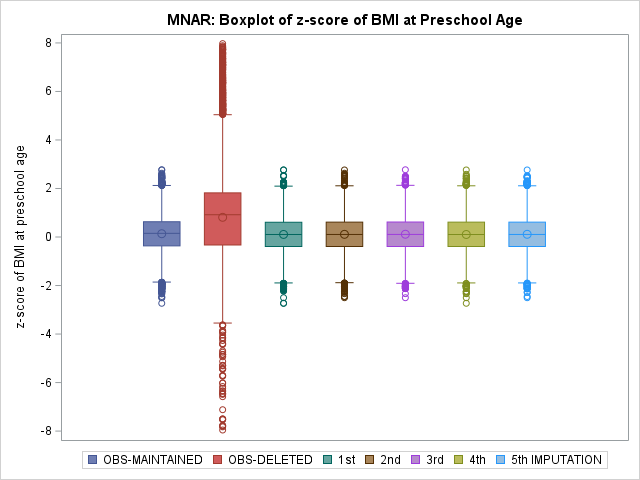

Note: BMI: body mass index; CIMD: Canadian Index of Multiple Deprivation; MAR: missing at random; MCAR: missing completely at random; MNAR: missing not at random; OBS: observed. Imputation model included the following variables: child age at BMI measurement, sex, birth weight, gestation, in-hospital birth, health zone, maternal age, marital status, parity, CIMD 2016 – Economic Dependency, CIMD 2016 – Situational Vulnerability, ethnicity, diabetes, hypertension, pre-eclampsia /eclampsia, cardiovascular and/or renal disease.
